# Frozen elephant trunk repair in heritable thoracic aortic disease: Impact of genetic aortopathy on long-term outcomes – A multicenter analysis

**DOI:** 10.64898/2026.06.09.26355316

**Authors:** Tim Berger, Sven Peterss, Leonard Pitts, Jörg Kempfert, Maria Nucera, Murat Yildiz, Tomas Holubec, Isabelle Haas, Martin Czerny, Maximilian Kreibich, Joseph Kletzer, Philipp Discher, Jenny Bialczak, Till Demal, Christian Detter, Simone Gasser, Maximilian Luehr, Anastasiia Alokhina, Konstantinos Tsagakis, Daniel Sebastian Dohle, Philipp Pfeiffer, Caroline Radner, Maximilian Pichlmaier, Nora Goebel, Bartosz Rylski, Zsuzsanna Arnold, Martin Grabenwoeger, Marie-Elisabeth Stelzmueller, Julia Dumfarth, Florian Schoenhoff, Jens Brickwedel

## Abstract

**Aims:** This multicenter study aims to compare outcomes of total aortic arch replacement (TAR) using the frozen elephant trunk (FET) technique in patients with and without heritable thoracic aortic disease (HTAD) and to assess whether HTAD influences postprocedural adverse aortic events (AAEs).

**Methods:** From 06/2007 to 05/2024, aortic databases from 13 European centers were screened for HTAD patients undergoing TAR with FET. All consecutive dissection and aneurysm non-HTAD patients from the four core centers served as comparator. The primary outcome was AAE, a composite of diameter progression, distal stent graft induced new entry (dSINE), malperfusion, rupture and pseudoaneurysm at 5 years after FET implantation.

**Results:** Of 2739 FET patients, 196 (7.2%) were diagnosed with HTAD. The control group consisted of 867 non-HTAD FET patients. Marfan syndrome was the most common condition (72%), followed by Loeys-Dietz syndrome (11%), vascular Ehlers-Danlos syndrome (5.6%) and Turner syndrome (2.0%). Seventeen (8.8%) patients were diagnosed with ns-HTAD. At 5 years 46 (24%) AAEs occurred in the HTAD group, 169 (20%) in the non-HTAD group (p=0.2). Diameter progression was the most common event (10% vs. 12%; p=0.6), followed by dSINE (5.8% vs. 4.5%; p=0.5), malperfusion (4.2% vs. 3.3%; p=0.5), rupture (2.1% vs. 0.7%; p=0.09) and pseudoaneurysm (0.5% vs. 0.2%; p=0.5).

**Conclusions:** The FET technique appears safe and effective for acute and chronic aortic disease in HTAD patients, with outcomes comparable to non-HTAD cases and no increase in graft-related complications, challenging traditional concerns about stent graft use in genetically mediated aortic disease.

**Graphical Element:** 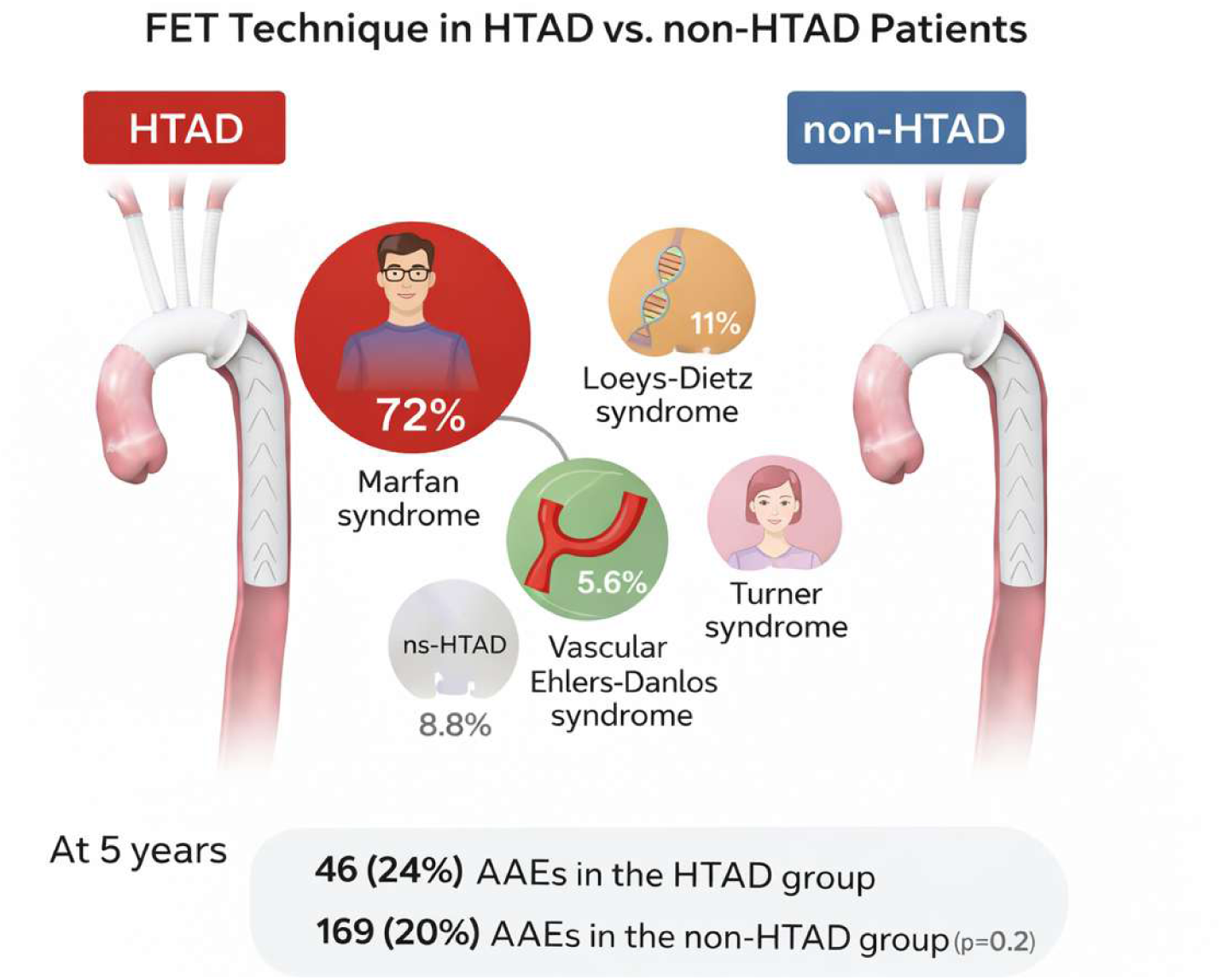

## Introduction

Heritable thoracic aortic diseases (HTAD), including Marfan-, Loeys–Dietz-, vascular Ehlers–Danlos and Turner syndrome, as well as non-syndromic forms, comprise a group of genetically mediated connective tissue disorders predisposing to early-onset thoracic aortic aneurysm, dissection, and rupture^1^. Current guidelines list 12 genes in which pathogenic variants have direct therapeutic consequences. Advances in diagnosis and surgical management have improved life expectancy of patients with heritable aortopathy; however, progressive distal aortic disease often necessitates complex reinterventions^2^. Repair of extensive arch and descending aortic pathology remains particularly challenging in these patients due to the fragility and altered biomechanics of the aortic wall.

The frozen elephant trunk (FET) technique combines open arch replacement with antegrade stent graft deployment into the descending aorta, enabling single-stage repair and providing a durable landing zone for future interventions^3,4^. Its efficacy and safety have been established in large series of patients with degenerative and post-dissection aortic disease^5–7^. However, the use of a stent graft in weakened aortic tissue in heritable aortopathies remains controversial. Concerns persist regarding stent-induced injury, distal aneurysm formation, and long-term durability in HTAD patients. Consequently, major guidelines advise caution or avoidance of endovascular repair in connective-tissue disorders, except in emergency settings^8^.

Nonetheless, the FET technique differs fundamentally from conventional thoracic endovascular aortic repair (TEVAR). It is a hybrid graft consisting of a proximal dacron and distal stent graft part. The hybrid graft is deployed under direct vision while the aortic arch is replaced surgically, potentially reducing mechanical stress on diseased tissue and eliminating the need for a proximal sealing zone. Evidence supporting or refuting its use in HTAD is limited, with most studies restricted to small, single-center Marfan cohorts^9,10^.

This multicenter study aims to compare outcomes of total aortic arch replacement (TAR) using the FET technique between patients with and without HTAD. By analyzing a large cohort across specialized aortic centers, we aimed to determine whether HTAD is associated with increased adverse aortic events, reintervention, or mortality, and to evaluate the safety and durability of FET in genetically mediated aortic disease.

## Patients and methods

### Ethical statement

IRB approval for this multicenter study was obtained on 30/07/2024 (24-1247-S1-retro) from the Ethics Committee of the University of Freiburg and the need for individual informed consent was waived due to the retrospective design. Anonymous data sharing was approved at each individual participating center.

### Data Collection Instruments

Between 06/2007 and 05/2024, the institutional aortic databases of 13 European tertiary aortic centers were screened for patients who underwent TAR using the FET technique. Subsequently, all individual patient records were reviewed, and patients with HTAD were identified. Baseline characteristics, aortic anatomy, intraoperative details, as well as postoperative and long-term outcomes, were assessed. Follow-up was conducted at the discretion of each participating center, and data were obtained from medical records or national/federal databases. The participating centers and their respective patient numbers are listed in **Supplemental Table 1.**

The study group comprised all HTAD patients (diagnosed based on the Ghent criteria or confirmed by genetic testing) treated at the 13 European aortic centers. All consecutive non-HTAD patients with dissection or aneurysm treated during the same period at four core centers (Bern, Freiburg, Hamburg, and Munich) served as the comparator group for the analysis of the primary outcome. The non-HTAD group comprises all patient in whom no clinical or genetic diagnosis of HTAD has been established.

### Primary Outcome

The primary outcome was adverse aortic event, a composite outcome measure of diameter progression, distal stent graft induced new entry (dSINE), malperfusion, rupture and pseudoaneurysm at 5 years after FET implantation in HTAD patients compared to non-HTAD patients.

### Secondary Outcomes

Secondary outcomes include the individual items of the primary outcome as well as the occurrence rates of aortic reintervention and mortality at 5 years. Additional secondary 30-day outcomes include the rates of the following parameters: mortality, stroke, paraplegia, renal replacement therapy, tracheostomy, duration of hospital and intensive care unit stay.

### Definition of Parameters

HTAD was subclassified in syndromic and non-syndromic HTAD. The syndromic subgroup included Marfan syndrome, Loeys-Dietz syndrome, Turner syndrome, vascular Ehlers-Danlos syndrome and osteogenesis imperfect. Non-syndromic HTADs include patients with pathogenic variants in ACTA2, MYH11, MYLK, LOX and PRKG1. HTAD was diagnosed based on clinical evaluation and genetic testing ^8,11–13^.

Adverse aortic events were assessed during follow-up, with only the first event considered in cases of multiple occurrences. Mortality was defined as death from any cause. Stroke was defined as a clinically evident neurological deficit accompanied by radiological confirmation of ischemia or hemorrhage. Aortic diameter progression was defined as an increase in aortic diameter of ≥0.5 cm per year^8^. The TEM classification was applied to assess type, entry, and malperfusion of acute aortic dissection^14^. The GERAADA score was calculated to anticipated 30 days mortality in patients with acute type A aortic dissection^15^. Emergent procedures were defined as those performed within 24 hours, while urgent procedures were conducted during the same hospital stay as the index event.

### Genetic Testing

Genetic testing was performed according to current guidelines in thoracic aortic disease patients <60 years of age, patients with a family history of thoracic aortic aneurysm or dissection, patients with syndromic features and those with more than one affected aortic segment. Genetic testing was performed using commercially available platforms. Number and type of genes varied across different centers, as well as over time. All panels included those genes where pathogenic variants carry therapeutic consequences in current guidelines (FBN1, TGFBR1, TGFBR2, SMAD3, SMAD2, TGFB2, TGFB3, COL3A1, PRKG1, MYLK, MYH11 and ACTA2). Classification of variants was established using 2015 ACMG criteria. Only variants considered pathogenic or likely-pathogenic were considered for this study.

### Statistical analysis

Statistical analyses were conducted using R (version 4.3.3) within the RStudio environment. Data visualizations were produced with the ggplot2 package (version 3.5.0). The Shapiro–Wilk test was employed to assess the normality of data distributions. Variables following a normal distribution are presented as means with standard deviations and were compared using unpaired t-tests. Non-normally distributed variables are reported as medians with interquartile ranges and analyzed using Mann–Whitney U tests. Categorical variables were evaluated using the Chi-squared test or, where appropriate, Fisher’s exact test.

Survival probabilities were estimated using the Kaplan-Meier Survival method. Group comparisons were performed with the log-rank test, and survival curves were plotted to illustrate time-to-event distributions. Patients were censored at last known follow-up if no event occurred.

Cumulative incidence functions for adverse events and death were calculated to account for competing risks. The Fine and Gray subdistribution hazards model was employed for multivariable analyses, allowing estimation of the effect of covariates on cumulative incidence in the presence of competing risks. Cumulative incidence curves were generated accordingly.

Interaction analyses were conducted by incorporating multiplicative interaction terms between the primary exposure and prespecified covariates within the regression models (Cox proportional hazards or Fine and Gray subdistribution hazards models, as appropriate). The validity of the proportional hazard’s assumption was assessed using Schoenfeld residuals, while the linearity and model fit were examined through residual diagnostics. The statistical significance of interaction effects was tested using Wald or likelihood ratio tests, and results were further explored via subgroup analyses and interaction plots to clarify the nature and direction of modification.

An alpha value of 0.05 was set to determine statistical significance.

## Results

### Patient characteristics

Out of 2739 patients treated at the 13 European aortic centers, 196 (7.2%) were clinically or genetically diagnosed with HTAD. The control group of the four core centers consisted of 867 FET patients treated for aortic dissection or aneurysm. Age (47.00 (36.00, 54.75) HTAD vs. 66.00 (58.64, 73.00) years non-HTAD, p<0.001) and sex (female - 85 (43%)) HTAD vs. 298 (34%) non-HTAD; p=0.018) differed significantly between HTAD and non-HTAD patients. Cardiovascular risk factors were more common in the non-HTAD group. Preoperative characteristics and comorbidities are summarized in **Table 1**.

**Table 1.** Patient characteristics.

| Characteristic | Overall N = 1,063<sup>1</sup> | HTAD Status |  | p-value<sup>2</sup> |
| --- | --- | --- | --- | --- |
|  |  | HTAD N = 196<sup>1</sup> | Non-HTAD N = 867<sup>1</sup> |  |
| Age (years) | 63.00 (53.00, 71.17) | 47.00 (36.00, 54.75) | 66.00 (58.64, 73.00) | <0.001 |
| Sex (female) | 383 (36%) | 85 (43%) | 298 (34%) | 0.018 |
| Height (cm) | 176.00 (168.00, 182.00) | 182.00 (173.00, 190.00) | 175.00 (168.00, 180.00) | <0.001 |
| Weight (kg) | 80.00 (69.00, 93.00) | 81.50 (68.00, 95.00) | 80.00 (70.00, 92.00) | 0.5 |
| BMI | 26.00 (23.14, 29.00) | 24.10 (21.55, 27.10) | 26.12 (23.51, 29.20) | <0.001 |
| Obesity | 247 (25%) | 27 (14%) | 220 (28%) | <0.001 |
| Dyslipidemia | 388 (37%) | 32 (16%) | 356 (42%) | <0.001 |
| History of Hypertension | 841 (80%) | 125 (64%) | 716 (83%) | <0.001 |
| History of smoking |  |  |  | <0.001 |
| Active Smoker | 255 (24%) | 23 (12%) | 232 (27%) |  |
| Non Smoker | 657 (63%) | 160 (83%) | 497 (59%) |  |
| Past Smoker | 130 (12%) | 10 (5.2%) | 120 (14%) |  |
| Diabetes Mellitus |  |  |  | 0.019 |
| Insulin dependent Diabetes Mellitus | 28 (2.7%) | 3 (1.5%) | 25 (2.9%) |  |
| No Diabetes Mellitus | 999 (95%) | 193 (98%) | 806 (94%) |  |
| Non-Insulin dependent Diabetes Mellitus | 28 (2.7%) | 0 (0%) | 28 (3.3%) |  |
| Renal failure |  |  |  | <0.001 |
| No Renal Failure | 891 (84%) | 183 (93%) | 708 (82%) |  |
| Renal Failure (Dialysis) | 41 (3.9%) | 2 (1.0%) | 39 (4.5%) |  |
| Renal Failure (No Dialysis) | 125 (12%) | 11 (5.6%) | 114 (13%) |  |
| Bicuspid aortic valve | 31 (2.9%) | 4 (2.1%) | 27 (3.1%) | 0.4 |
| Previous stroke | 78 (9.1%) | 17 (8.7%) | 61 (9.2%) | 0.8 |
| Marfan syndrome | 141 (13%) | 141 (47.6%) | 0 (0%) | <0.001 |
| Ehlers-Danlos syndrome | 11 (1.0%) | 11 (3.7%) | 0 (0%) | <0.001 |
| Loeys-Dietz syndrome | 22 (2.1%) | 22 (11.2%) | 0 (0%) | <0.001 |
| Non-syndromatic TAAD | 17 (1.8%) | 17 (8.7%) | 0 (0%) | <0.001 |
| Turner Syndrome | 4 (0.4%) | 4 (2.0%) | 0 (0%) | 0.001 |
| Previous Cardiac/Aortic Surgery | 402 (38%) | 131 (67%) | 271 (31%) | <0.001 |
| Previous aortic valve replacement | 62 (5.8%) | 27 (14%) | 35 (4.0%) | <0.001 |
| Previous valve sparing root replacement | 51 (4.8%) | 32 (16%) | 19 (2.2%) | <0.001 |
| Previous valve carrying conduit | 95 (8.9%) | 57 (29%) | 38 (4.4%) | <0.001 |
| Previous supracoronary ascending replacement | 230 (22%) | 61 (31%) | 169 (19%) | <0.001 |
| Previous TEVAR | 60 (5.6%) | 19 (9.7%) | 41 (4.7%) | 0.007 |

<sup>1</sup>Median (Q1, Q3); n (%)
<sup>2</sup>Wilcoxon rank sum test; Pearson's Chi-squared test; Fisher's exact test

### Heritable thoracic aortic disease characteristics

Genetic testing data were available from all centers. In these centers, a total of 2739 patients underwent treatment with the FET technique. Among them, 223 (8.1%) patients received genetic testing, of whom 148 (5.4%) were identified as carriers of a pathogenic variant. In 128 (4.7) patients, the diagnosis of HTAD was already established before the time of the FET procedure.

Marfan syndrome was the most common HTAD condition (n=141, 72%), followed by Loeys-Dietz syndrome (n=22, 11%), vascular Ehlers-Danlos syndrome (n=11; 5.6%) and Turner syndrome (n=4; 2.0%). Seventeen (8.8%) patients were diagnosed with ns-HTAD.

Previous aortic procedures were common in both groups with a significant higher rate in the HTAD group. The HTAD group consisted of more patients with aortic dissection, whereas there was no difference in the type of acute aortic dissection based on the TEM classification between HTAD and non-HTAD patients. Patients with HTAD were more frequently treated for residual aortic dissection after previous repair for acute type A aortic dissection. Aortic characteristics are listed in **Table 2**.

**Table 2.** Aortic disease characteristics at the time of the frozen elephant trunk procedure.

| Characteristic | Overall N = 1,063<sup>1</sup> | HTAD Status |  | p-value<sup>2</sup> |
| --- | --- | --- | --- | --- |
|  |  | HTAD N = 196<sup>1</sup> | Non-HTAD N = 867<sup>1</sup> |  |
| Acute (1-14 days) | 375 (35%) | 71 (36%) | 304 (35%) | 0.8 |
| Subacute (15-90 days) | 30 (2.8%) | 10 (5.1%) | 20 (2.3%) | 0.033 |
| Chronic (>90 days) | 482 (45%) | 101 (52%) | 381 (44%) | 0.056 |
| Aortic dissection | 667 (63%) | 162 (83%) | 505 (59%) | <0.001 |
| Intramural hematoma | 20 (1.9%) | 3 (1.5%) | 17 (2.0%) | >0.9 |
| Penetrating aortic ulcer | 12 (1.1%) | 1 (0.5%) | 11 (1.3%) | 0.7 |
| Type of Aortic Dissection |  |  |  | 0.9 |
| Type A | 337 (60%) | 89 (60%) | 248 (59%) |  |
| Type B | 161 (28%) | 40 (27%) | 121 (29%) |  |
| Type None-A None-B | 67 (12%) | 19 (13%) | 48 (12%) |  |
| Primary aneurysm | 324 (31%) | 16 (8.2%) | 308 (36%) | <0.001 |
| Post-dissection aneurysm | 230 (22%) | 88 (45%) | 142 (16%) | <0.001 |

<sup>1</sup>n (%)<sup>2</sup>Pearson's Chi-squared test; Fisher's exact test**Table 3. Outcomes**

### Intraoperative data

There were more urgent procedures in the HTAD group (n=26, 13% HTAD vs. n=52, 6% non-HTAD). The Thoraflex (Terumo Aortic, Inchinnan UK) prothesis was used in 130 (66%) HTAD patients, whereas the Evita (Artivion, Kennesaw, GA, USA) was implanted in 64 (33%) patients. Intraoperative details including landing zone, canulation site, concomitant procedures and intraoperative times are listed in **Supplemental Table 2.**

### Primary outcome – adverse aortic events

At 5 years 46 (24%) adverse aortic events occurred in the HTAD group, 169 (20%) in the non-HTAD group (p=0.2). Diameter progression was the most common event (n=20, 10% HTAD vs. n=102, 12% non- HTAD; p=0.6), followed by dSINE (n=11, 5.8% HTAD vs. n=39, 4.5% non- HTAD; p=0.5), malperfusion (n=8, 4.2% HTAD vs. n=28, 3.3% non- HTAD; p=0.5), rupture (n=4, 2.1% HTAD vs. n=6, 0.7% non- HTAD; p=0.09) and pseudoaneurysm (n=1, 0.5% HTAD vs. n=2, 0.2% non- HTAD; p=0.5). The cumulative incidence curve with competing risk of death for the primary outcome is illustrated in **Figure 1**.

**Figure 1.**
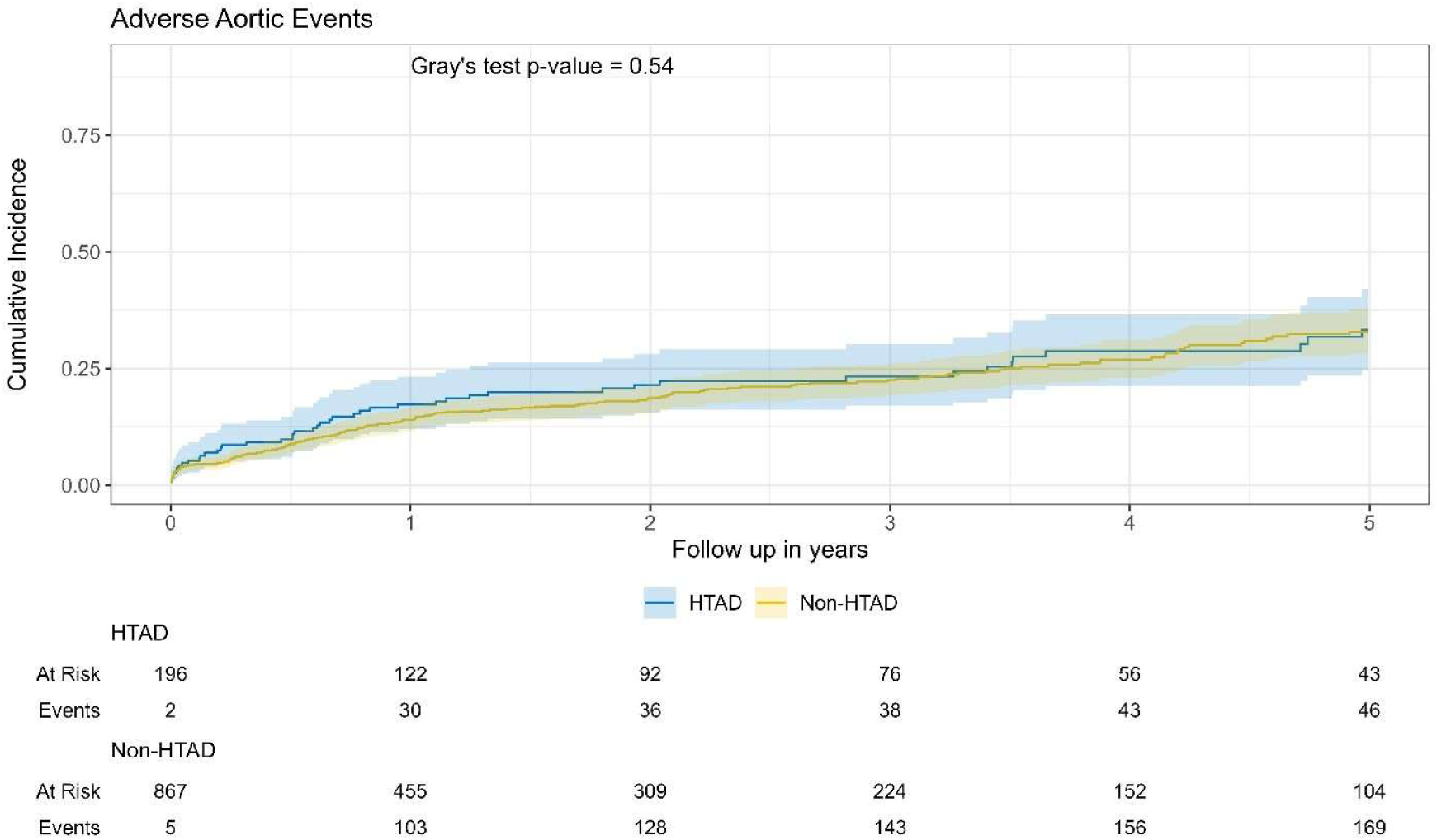
Cumulative incidence of adverse aortic events in heritable thoracic aortic disease patients (HTAD) and patients without heritable thoracic aortic disease (Non-HTAD).

The cumulative incidence of adverse aortic events was comparable in all HTAD subgroups, including Marfan syndrome, Loeys-Dietz syndrome, Turner syndrome, vascular Ehlers-Danlos syndrome and nsHTAD (**Figure 2**).

**Figure 2.**
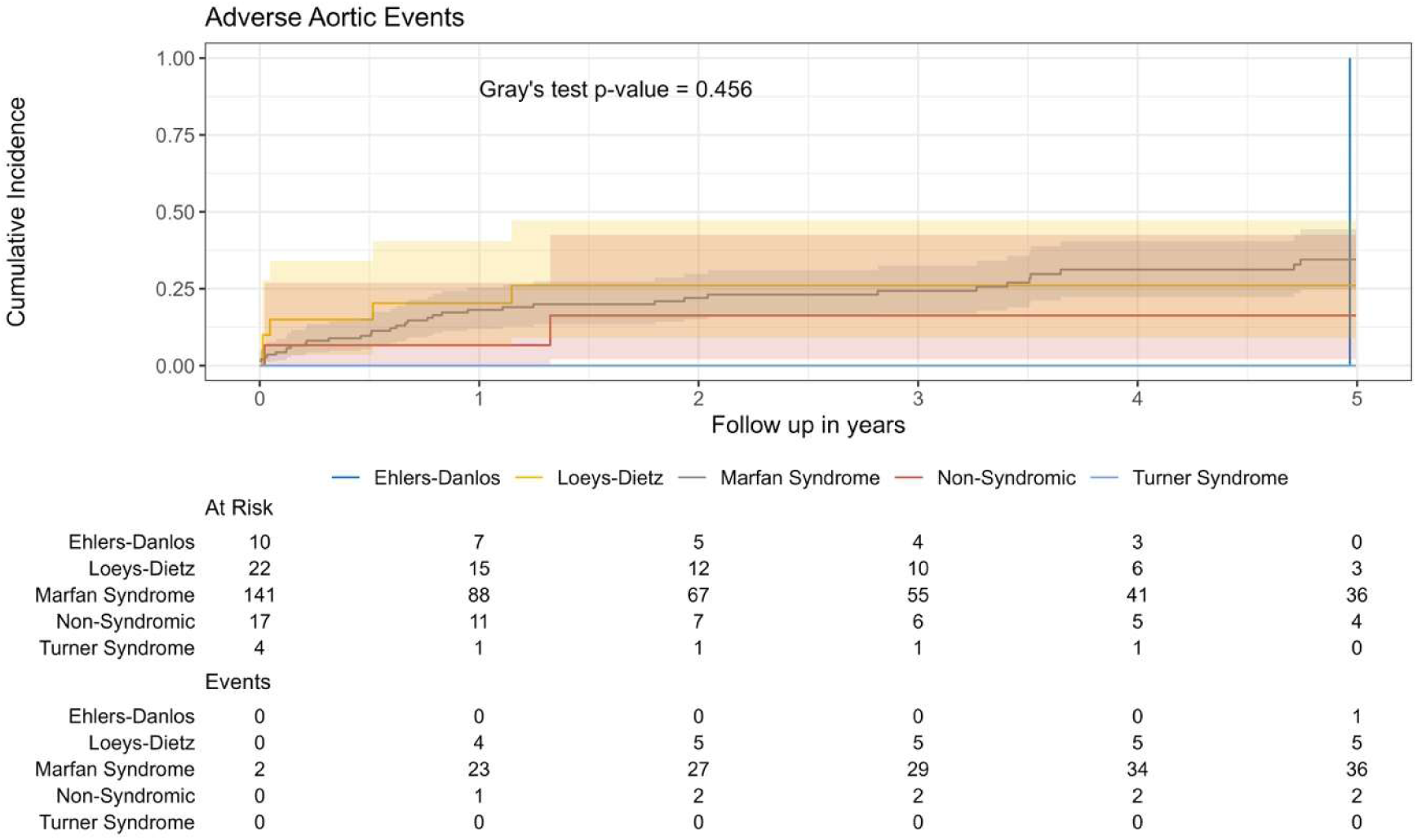
Cumulative incidence of adverse aortic events patients with Ehlers-Danlos, Loeys-Dietz syndrome, Marfan syndrome, non-syndromic and Turner syndrome.

### Secondary outcomes

30-day mortality (n=6, 3.1% HTAD vs. n=106, 12% non-HTAD; p<0.001) and postoperative stroke (n=13, 6.6% HTAD vs. n=105, 14% non-HTAD; p<0.001) was significantly lower in the HTAD group. Other secondary outcomes including bleeding, paraplegia, tracheostomy and dialysis were comparable as well as in-hospital and ICU stay. Outcomes are summarized in **Table 3**.

**Table 3.**
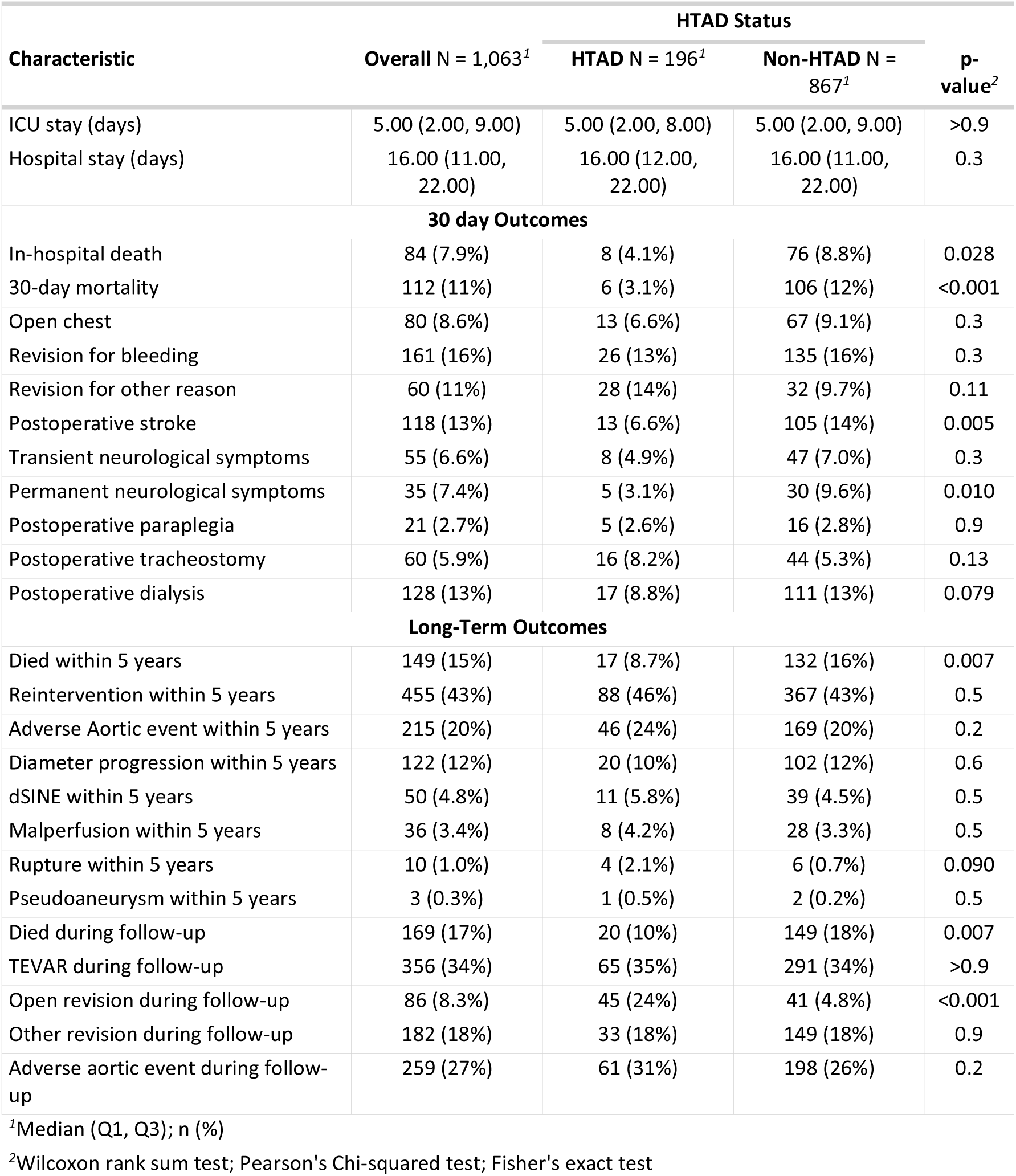
Outcomes.

| Characteristic | Overall N = 1,063<sup>1</sup> | HTAD Status |  | p-value<sup>2</sup> |
| --- | --- | --- | --- | --- |
|  |  | HTAD N = 196<sup>1</sup> | Non-HTAD N = 867<sup>1</sup> |  |
| ICU stay (days) | 5.00 (2.00, 9.00) | 5.00 (2.00, 8.00) | 5.00 (2.00, 9.00) | >0.9 |
| Hospital stay (days) | 16.00 (11.00, 22.00) | 16.00 (12.00, 22.00) | 16.00 (11.00, 22.00) | 0.3 |
| **30 day Outcomes** |  |  |  |  |
| In-hospital death | 84 (7.9%) | 8 (4.1%) | 76 (8.8%) | 0.028 |
| 30-day mortality | 112 (11%) | 6 (3.1%) | 106 (12%) | <0.001 |
| Open chest | 80 (8.6%) | 13 (6.6%) | 67 (9.1%) | 0.3 |
| Revision for bleeding | 161 (16%) | 26 (13%) | 135 (16%) | 0.3 |
| Revision for other reason | 60 (11%) | 28 (14%) | 32 (9.7%) | 0.11 |
| Postoperative stroke | 118 (13%) | 13 (6.6%) | 105 (14%) | 0.005 |
| Transient neurological symptoms | 55 (6.6%) | 8 (4.9%) | 47 (7.0%) | 0.3 |
| Permanent neurological symptoms | 35 (7.4%) | 5 (3.1%) | 30 (9.6%) | 0.010 |
| Postoperative paraplegia | 21 (2.7%) | 5 (2.6%) | 16 (2.8%) | 0.9 |
| Postoperative tracheostomy | 60 (5.9%) | 16 (8.2%) | 44 (5.3%) | 0.13 |
| Postoperative dialysis | 128 (13%) | 17 (8.8%) | 111 (13%) | 0.079 |
| **Long-Term Outcomes** |  |  |  |  |
| Died within 5 years | 149 (15%) | 17 (8.7%) | 132 (16%) | 0.007 |
| Reintervention within 5 years | 455 (43%) | 88 (46%) | 367 (43%) | 0.5 |
| Adverse Aortic event within 5 years | 215 (20%) | 46 (24%) | 169 (20%) | 0.2 |
| Diameter progression within 5 years | 122 (12%) | 20 (10%) | 102 (12%) | 0.6 |
| dSINE within 5 years | 50 (4.8%) | 11 (5.8%) | 39 (4.5%) | 0.5 |
| Malperfusion within 5 years | 36 (3.4%) | 8 (4.2%) | 28 (3.3%) | 0.5 |
| Rupture within 5 years | 10 (1.0%) | 4 (2.1%) | 6 (0.7%) | 0.090 |
| Pseudoaneurysm within 5 years | 3 (0.3%) | 1 (0.5%) | 2 (0.2%) | 0.5 |
| Died during follow-up | 169 (17%) | 20 (10%) | 149 (18%) | 0.007 |
| TEVAR during follow-up | 356 (34%) | 65 (35%) | 291 (34%) | >0.9 |
| Open revision during follow-up | 86 (8.3%) | 45 (24%) | 41 (4.8%) | <0.001 |
| Other revision during follow-up | 182 (18%) | 33 (18%) | 149 (18%) | 0.9 |
| Adverse aortic event during follow-up | 259 (27%) | 61 (31%) | 198 (26%) | 0.2 |

<sup>1</sup>Median (Q1, Q3); n (%)
<sup>2</sup>Wilcoxon rank sum test; Pearson's Chi-squared test; Fisher's exact test

Five-year mortality was lower in the HTAD group (n=17, 8.7% HTAD vs. n=132, 16% non-HTAD; p=0.007), whereas reintervention rates were comparable (n=88, 46% HTAD vs. n=367, 43% non-HTAD; p=0.5)

### Subgroup analyses

Survival within five years postoperatively was comparable in patients with Marfan syndrome, Loeys-Dietz syndrome, Turner syndrome, vascular Ehlers-Danlos syndrome and nsHTAD (**Figure 3**).

**Figure 3.**
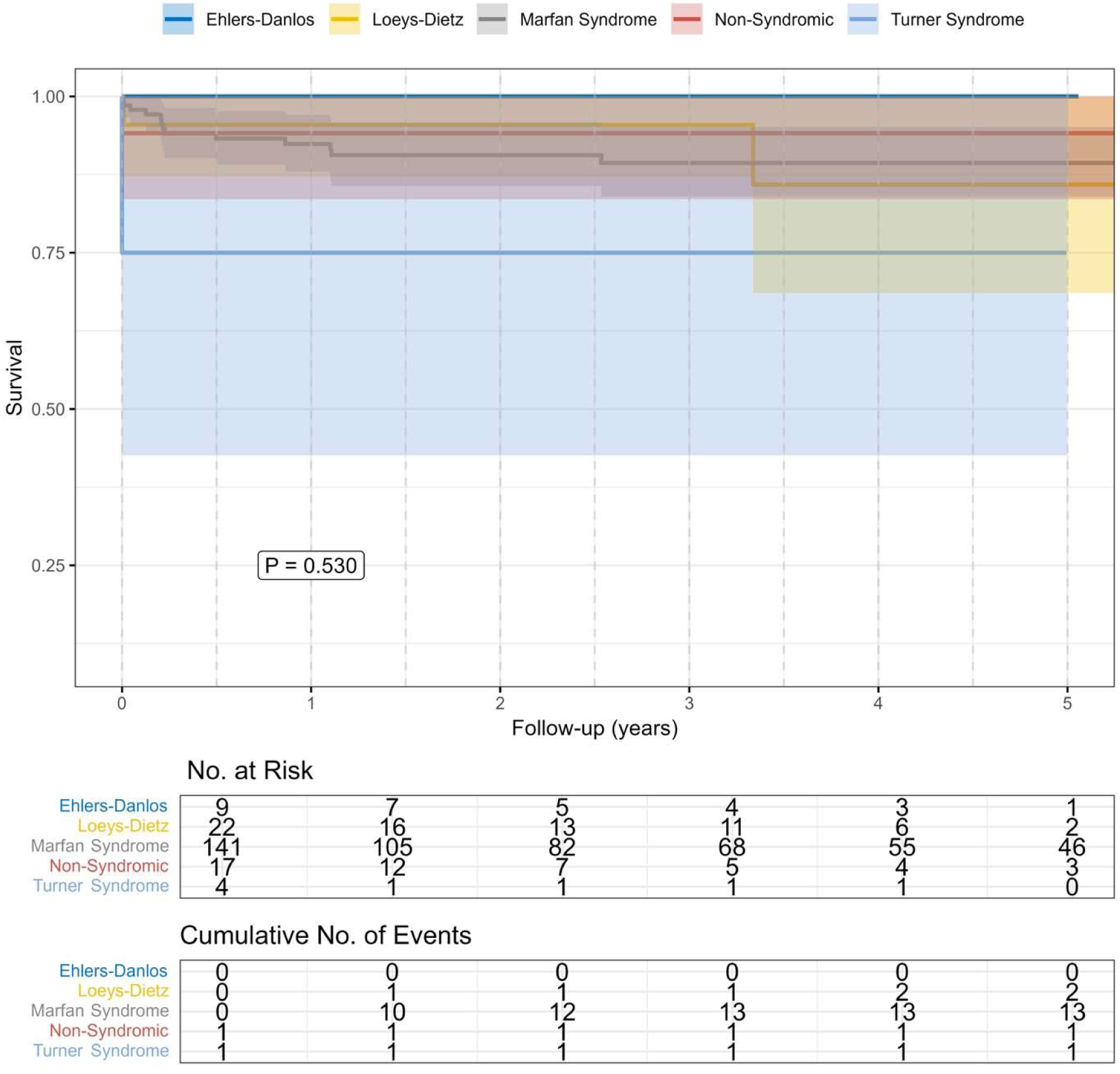
Kaplan-Meier estimates of survival for patients with Ehlers-Danlos, Loeys-Dietz syndrome, Marfan syndrome, non-syndromic and Turner syndrome.

The competing risk analysis (competing risk: death) revealed no differences regarding adverse aortic events and reinterventions in HTAD and non-HTAD patients.

There was no difference in the aortic dissection subgroup as well as in the aortic aneurysm subgroup (**Figure 4)** when comparing HTAD versus non-HTAD patients in terms of adverse aortic events and aortic reinterventions within 5 years postoperatively. Whereas HTAD patients with aortic dissection revealed a superior survival compared to non-HTAD patients at 5 years.

**Figure 4.**
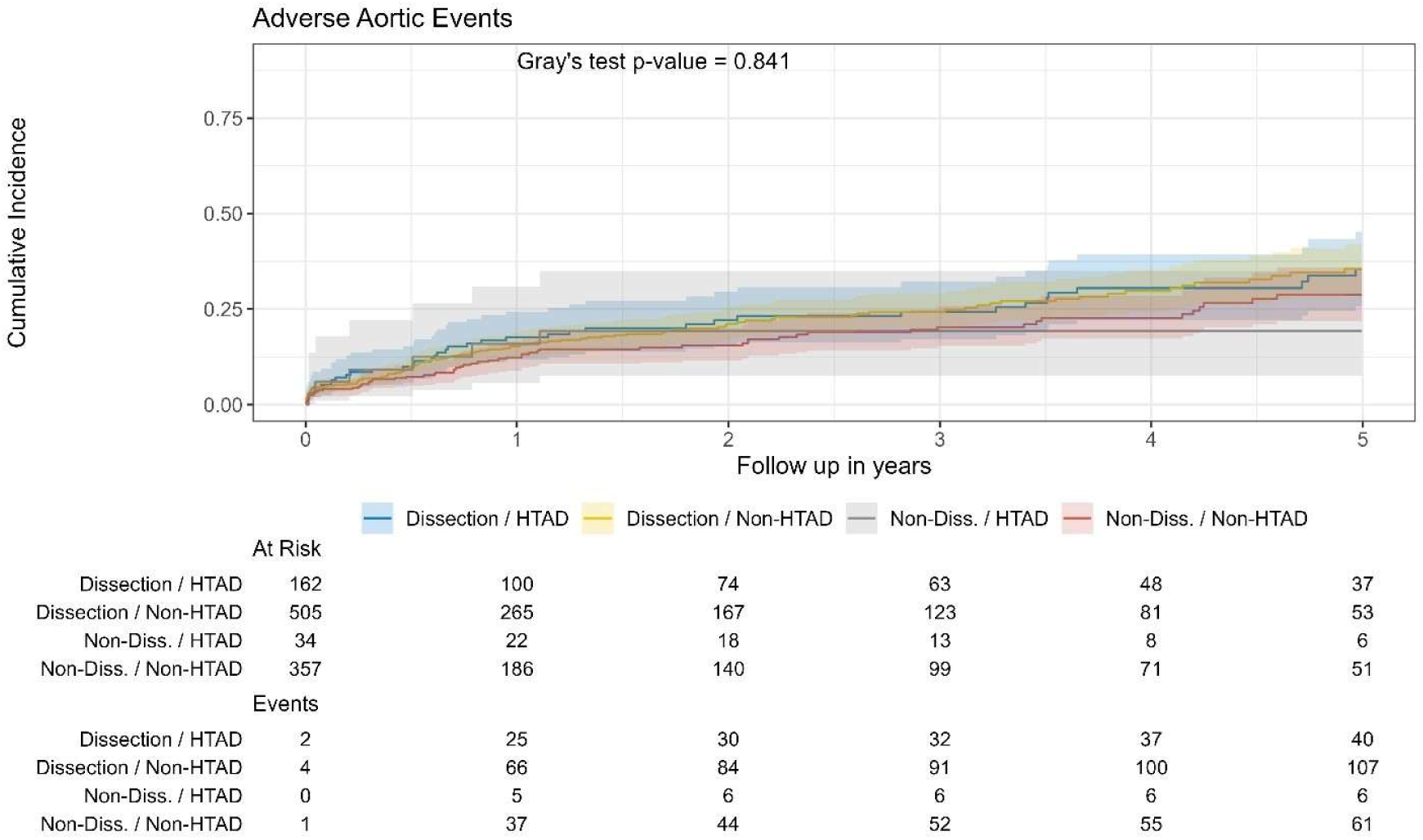
Cumulative incidence of adverse aortic events in dissection heritable thoracic aortic disease patients (Dissection / HTAD), in dissection non-heritable thoracic aortic disease patients (Dissection /Non-HTAD), in non-dissection heritable thoracic aortic disease patients (Non-Diss. / HTAD), in non-dissection non-heritable thoracic aortic disease patients (Non-Diss./Non-HTAD).

Comparative hazard ratio analyses identified significant subgroup-specific risk modulation between HTAD and non-HTAD cohorts, with several variables, such as male sex (interaction P = 0.006), dyslipidemia (interaction P = 0.007), and prior valve-carrying conduit (interaction P = 0.013), demonstrating meaningful effect heterogeneity in the mortality model. For adverse aortic events, significant interaction was observed for ascending aortic cannulation (interaction P = 0.006) and right axillary cannulation (interaction P = 0.026). However, after applying Bonferroni adjustment for multiple comparisons (33 factors: adjusted threshold p < 0.0015 for death, p < 0.0013 for adverse events), none of these interactions remained statistically significant, indicating that observed effects may be attributable to chance when controlling the family-wise error rate. A detailed graphical summary is provided in **Figure 5**.

**Figure 5.**
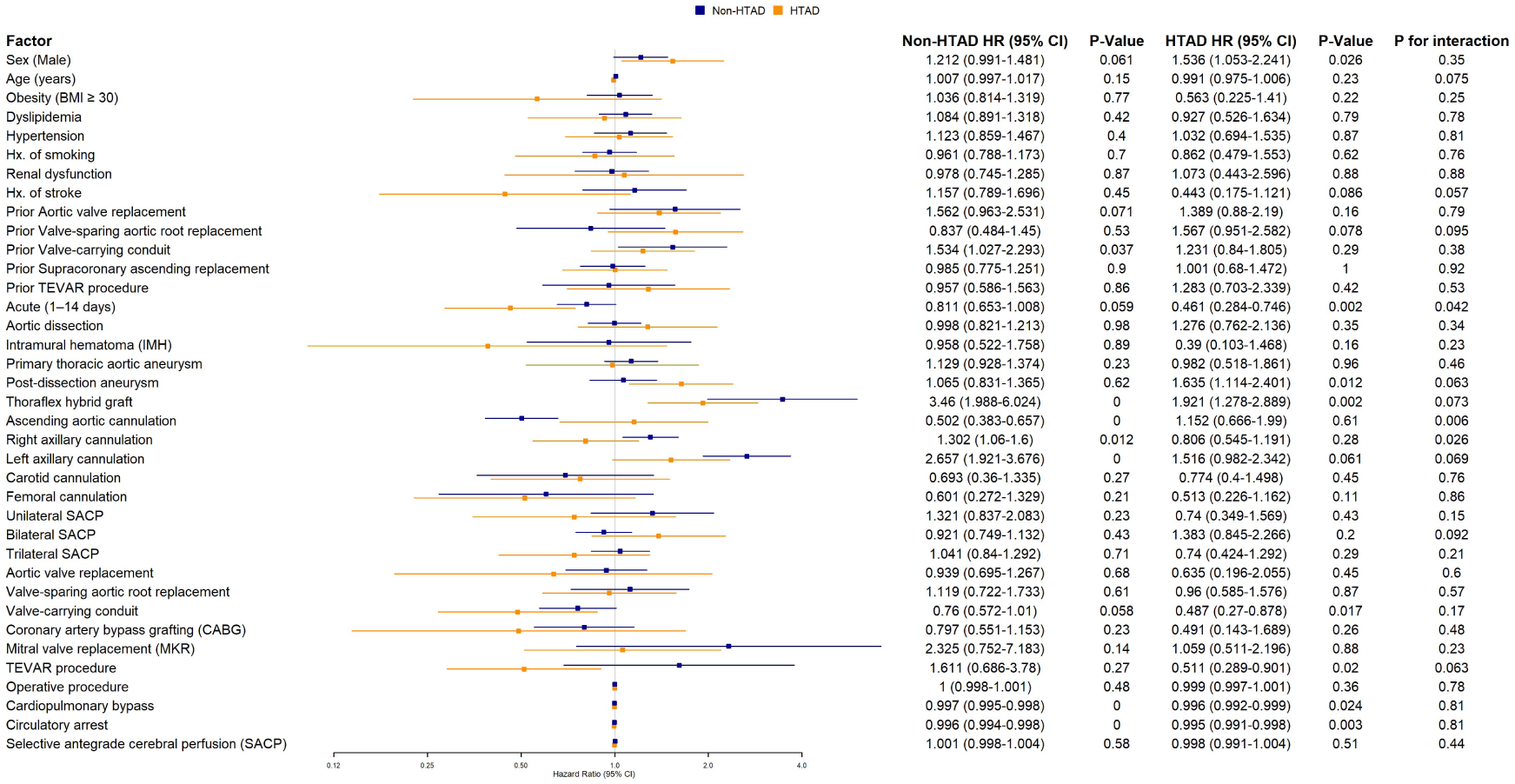
Interaction analysis of potential risk factors of adverse aortic events.

## Discussion

The most relevant findings and implications of this study can be summarized as follows: **I.** TAR replacement using the FET technique revealed comparable adverse aortic event rates in patients with HTAD compared to non-HTAD patients. **II.** There was no difference in adverse aortic events between the syndromic and non-syndromic HTAD subgroups. **III.** Therefore, the FET technique is encouraged for the treatment of acute and chronic aortic pathologies in patients with HTAD.

The baseline characteristics of the study cohort were largely comparable to previously published reports on patients with HTAD^16,17^. The distribution of demographic and clinical variables within the HTAD group closely reflects those described in other large single- and multicenter experiences focusing on Marfan syndrome and related disorders^9,17,18^. Similarly, the overall characteristics of the entire study population, including patients without HTAD, align well with data from major multicenter registries evaluating TAR using the FET technique^5–7^. As anticipated, a substantial difference between the HTAD and non-HTAD groups was observed, as patients with HTAD were significantly younger and exhibited fewer traditional cardiovascular risk factors such as hypertension, diabetes, and atherosclerosis. Interestingly, there was no difference in acute type B aortic dissections in both groups. Adjustment for baseline characteristics was not performed, as these two populations represent inherently different disease entities rather than comparable subgroups within the same pathological spectrum. This fundamental biological difference cannot be fully addressed by statistical matching or regression techniques. Nevertheless, the focus of this study was not on comparing comorbidities but rather on evaluating the durability and interaction of the FET prosthesis with the underlying aortic tissue. Therefore, the distinct baseline profile of HTAD patients does not confound the principal findings, which concern long-term outcomes and prosthesis–aorta interaction in genetically weakened aortas compared with the relatively more resilient non-HTAD aortas.

Patients with HTAD exhibit intrinsic structural weakness of the aortic wall due to altered extracellular matrix composition, reduced elastin integrity, and increased collagen deposition, resulting in higher aortic stiffness and lower compliance. These pathophysiologic alterations impair the normal adaptive remodeling response to hemodynamic stress, as the aortic wall cannot thicken adequately to counteract circumferential stress in accordance with the Law of Laplace^1^. Consequently, the implantation of a stent graft with substantial radial force introduces additional biomechanical stress at the interface between the prosthesis and the fragile native aorta. In contrast to degenerative or atherosclerotic aortas, HTAD aortas are less capable of dissipating this radial load, raising theoretical concerns of graft-induced intimal injury, distal stent-induced new entry, or progressive dilation adjacent to the landing zones^19^. Nonetheless, recent clinical data, including our multicenter findings, suggest that when applied in specialized centers with careful patient selection and tailored device sizing, the FET technique does not lead to excess graft-related complications in HTAD patients. This observation challenges the traditional hesitancy toward hybrid approaches in genetically mediated aortopathies. Ultimately, these results indicate that while the biomechanical fragility of HTAD aortas warrants respect, it does not necessarily preclude the safe and durable application of modern stent graft technologies, especially when the proximal part is concomitantly replaced with a dacron graft.

The distribution of HTAD conditions mirrors the relative prevalence of these disorders reported in contemporary genetic and surgical series. Studies focusing exclusively on Marfan patients undergoing FET repair have previously shown encouraging outcomes regarding long-term survival and freedom from reintervention^10^. However, data on non-Marfan HTAD conditions remain limited. Our findings contribute to the growing evidence that the FET procedure can be safely applied across the spectrum of HTAD subtypes, including Loeys–Dietz and Ehlers–Danlos syndromes, provided that surgical indications and technical nuances are carefully adapted, which is still under debate for TEVAR.

Concomitant procedures were frequent in both groups, particularly aortic root replacement, which was performed more often among HTAD patients. This finding is consistent with the high prevalence of root aneurysms and connective tissue fragility in this subgroup. Previous studies have demonstrated that concomitant root replacement does not increase perioperative risk in patients undergoing TAR, and our results support this notion^20^. In our cohort, postoperative outcomes—including mortality and major complications—were at least comparable, and in some measures even superior, in the HTAD group compared with non-HTAD patients.

Despite these overall similarities, certain procedure-specific differences were noted, including variation in prosthesis types, cannulation sites, and distal landing zones. These differences likely reflect center-specific preferences and expertise rather than disease-specific technical requirements^21–23^. It is important to interpret these findings within the context of the study design: the HTAD group was derived from 13 specialized centers, whereas the control group originated from four core centers, which introduces an element of inter-center heterogeneity. Such variations are a known limitation of multicenter analyses and may affect operative details more than postoperative outcomes.

Adverse aortic event rates were comparable between HTAD and non-HTAD patients, both for the composite endpoint and for individual components. Remarkably, no significant difference was observed between the syndromic and non-syndromic HTAD subgroups, suggesting that the FET procedure is equally safe and effective across different genetic backgrounds. The safety of stent graft implantation remains controversial, especially in TEVAR, where two non-diseased affected landing zones are required. While TEVAR offers a less invasive alternative and may serve as a life-saving or bridging procedure in acute or complex scenarios, concerns persist regarding its long-term durability in genetically weakened aortic tissue. The fragile aortic wall in HTAD increases the risk of stent-induced injury, new dissection, and distal disease progression, leading to higher reintervention rates compared with non-HTAD patients. Current evidence is limited to small series and registry data, and guidelines as well as consensus documents recommend restricting TEVAR to highly selected cases or emergency situations when open repair is not feasible^8^. Consequently, careful patient selection, gentle device sizing, and lifelong imaging surveillance are essential when considering TEVAR in this challenging population is advocated^11,12^. Our study revealed that these concerns using TEVAR are not applicable to the FET technique, that was able to offer durable results as the proximal sealing zone is replaced in an open fashion and fixed to the stent graft part, eliminating the risk of retrograde dissection, stent graft migration. Of note, the proximal landing zone is crucial in the decision-making between open replacement using the FET technique or endovascular treatment.

The rate of aortic reintervention in the entire cohort was substantial yet consistent with other large studies assessing the FET technique^3,24,25^. Importantly, the incidence of reintervention did not differ between HTAD and non-HTAD patients, indicating that the interaction between the stent graft and the inherently weaker HTAD aortic wall does not translate into an increased risk of device-related failure or distal disease progression. Of note, despite substantial group differences, aortic pathologies were comparable, strengthening the durable treatment effect of the FET technique across all pathologies.

The observed differences in long-term mortality likely reflect the younger age and better overall health of HTAD patients, who are less burdened by cardiovascular comorbidities and exhibit superior perioperative resilience. This favorable baseline profile, rather than any procedural advantage, likely accounts for their relatively lower long-term mortality and morbidity. Nevertheless, the comparable event rates underscore the durability and safety of the FET approach in genetically mediated aortic disease, a key finding that supports its broader application in this challenging patient population.

## Limitations

This is a retrospective study with all limitations inherent to the study design. Patients were classified as having HTAD if a diagnosis had been previously established, based on the revised Ghent criteria or confirmed by genetic testing. However, not all patients undergoing FET received genetic testing, as achieving a 100% testing rate is challenging in a multicenter setting. Moreover, testing criteria and genes recommended for testing changed during the study period. Consequently, it cannot be entirely excluded that some patients with undiagnosed HTAD were included in the non-HTAD control group. Nevertheless, this potential overlap is unlikely to have affected the validity of our findings or the overall implications of the study.

## Conclusion

The present study reinforces the role of the frozen elephant trunk technique as a reliable and durable strategy for managing both acute and chronic aortic pathologies in patients with and without HTAD. The comparable outcomes between these groups, despite the inherent differences in age, tissue quality, and comorbidities, suggest that the FET technique can safely bridge the gap between emergency and elective treatment in complex aortic disease. Furthermore, the absence of excess graft-related complications in HTAD aortas challenges the traditional notion that stent graft implantation should be avoided in genetically mediated aortic disease.

## Acknowledgements

none

## Author contribution

Conceptualization: T.B., S.P., F.S., J.B. Data curation: J.K., T.B., P.D., P.P., M.N. C.R. K.T. L.P.; Z.A., J.D., J.B., M.L.,N.G., A.A., I.H., M.S., S.G., Formal Analysis: J.K., T.B.; Methodology: J.K., T.B.; Project administration: T.B.; Supervision: M.C., T.B.; Validation: J.K., T.B.; Visualization: J.K.; T.B.; Writing – original draft: J.K, T.B; Writing – review & editing: M.K., M.C, M.Y., M.G., M.L., P.D., P.P., M.N. C.R. K.T. L.P.; Z.A., J.D., J.B., N.G., B.R., D-S. D., S.P., F.S., J.B., I.H., T.H, M.S., S.G.,

## Data availability statement

The datasets analyzed during the current study are not publicly available due to strict institutional and ethical restrictions regarding patient data protection and confidentiality and are therefore not available for sharing. Data are available from the corresponding author on reasonable request, subject to approval by the relevant institutional review boards and data-sharing agreements with participating centers.

## Pre-registered clinical trial number

FRKS005257

